# Brief Multimodal Intervention to Address Bedtime Procrastination and Sleep through Self-Compassion and Sleep Hygiene during Stressful Times

**DOI:** 10.1101/2023.04.16.23288655

**Authors:** Steven L. Bistricky, Alicia K. Lopez, Tarryn B. Pollard, Alana Egan, Malena Gimenez-Zapiola, Bailey Pascuzzi, Kenia M. Velasquez, Miana Graves

## Abstract

Bedtime procrastination is increasingly recognized as a widespread impediment to health-promoting sleep. Based on its potential malleability, bedtime procrastination is starting to be targeted for intervention using traditional health behavior models, but other cognitive and emotional factors that potentially modulate bedtime procrastination warrant more targeted intervention. The present research recruited college students (n = 93) with self-reported tendencies toward bedtime procrastination and low self-compassion early in the COVID-19 pandemic, and it examined a hybrid intervention model involving a single group meeting and home practices that focused on comprehensive sleep hygiene or intentional self-compassion practices, simultaneously leveraging social motivation and commitment. It also examined bedtime procrastination, sleep, emotion regulation, and procrastinatory cognitions. The study showed evidence for feasibility, acceptability, reduced bedtime procrastination, improved sleep, and moderated mediation whereby the relationship between increased self-compassion and decreased bedtime procrastination was mediated by improved emotion regulation for those with elevated reductions in procrastinatory cognition. Predictors of bedtime procrastination reduction and other relevant sequelae differed between self-compassion and sleep hygiene virtual trainings. Thus, the present research expands and synthesizes a burgeoning literature, suggesting that integrating effective elements into acceptable interventions may help reverse a cycle of self-criticism, emotion dysregulation, bedtime procrastination, and sleep-related difficulties for many who might benefit.

## Introduction

Sleep insufficiency may cause or exacerbate a wide range of negative health outcomes, such as concentration and memory problems (1), mood and anxiety disorders, immunosuppression, cardiovascular disease, hypertension, obesity, diabetes, and increased cancer and morbidity risk (2–4). Sleep insufficiency is also among the most prominent causes of motor vehicle collisions, which are among the most common cause of preventable injury, psychiatric morbidity, and death (5,6).

More than one-third of adults in the U.S. regularly get insufficient sleep (Liu et al., 2016), and related concerns are driving demand for feasible, accessible, effective sleep promotion strategies (7). People get insufficient sleep for several reasons. Looking beyond deeply researched factors, there is growing attention in the research literature that some individuals regularly delay going to bed after their intended bedtime has passed. This sleep-reducing class of behavior is termed bedtime procrastination, the tendency to delay one’s intended bedtime without prevention from any external factors (8,9).

Even after accounting for well-known barriers to sleep, around half the population may go to bed later than planned three times a week or more (10,11). Importantly, this *bedtime procrastination* (BP) affects general adult populations whose sleep health is often not addressed in research or intervention development (9). BP is associated with measures of stress, anxiety, depression, well-being, as well as reduced sleep duration and sleep quality, and greater daytime fatigue (12,13). Further, BP partly mediates poor sleep outcomes (14–16). Thus, it is important to identify and target malleable factors that can reduce BP and address gaps in the literature (17,18).

BP is considered a maladaptive health behavior; whereas, going to bed at a relatively consistent set bedtime that allows for sufficient sleep duration would be a positive behavior to target for health benefits. The health belief model (HBM), theory of planned behavior (TPB), and the transtheoretical model of change (TTM) are among the most influential and well-supported theoretical frameworks that explain why people engage in adaptive or maladaptive health behaviors (e.g., vaccination uptake, physical exercise, diabetes management, sexually transmitted infection and cancer prevention, helping behavior, smoking, alcohol abuse, bedtime procrastination), and these models commonly inform interventions that effectively promote adaptive health behavior initiation and maintenance (19–27). According to the HBM, people more often engage in health behaviors if they 1) view practicing it as personally beneficial, doable, having few barriers, and important to valued health outcomes; 2) intend to engage in the behavior; and 3) are prompted to engage in the behavior (28–30). According to the TPB, having positive attitudes toward a health behavior, perceiving it as a desirable social norm to adhere to, and a belief one can effectively enact the behavior all increase planning and implementation of the behavior (31,32). The transtheoretical model (TTM) of behavior change also suggests that resolving ambivalence toward increased readiness for behavior change, planning behavior, and public commitment promotes completion of intended behaviors (33,34). Intervention factors, such as alliance with clients’ goals, can also help increase clients’ “change talk” about their desire, ability, reasons, and need to commit to change. Moving from commitment to change behavior is mediated by planning. Although all three models have been used to understand promotion of healthy sleep behaviors (26,32,35) they have thus far been only minimally applied to BP.

Suh and colleagues (36) recently piloted a TTM-inspired BP intervention in which participants completed three 50-minute weekly in-person individual sessions and an additional booster call. Participants who entered the study reporting high BP showed post-intervention improvements in BP, sleep efficiency, and sleep-related daytime fatigue. Investigating whether similar effects might be achieved with a lower treatment “dose,” Valshtein and colleagues (37) compared an ultra-low-provider-burden online intervention targeting health behavior model factors against two active control conditions. The intervention ultimately increased BP reduction commitment and behavior. Importantly, both interventions elicited large effect size reductions in BP. In sum, BP interventions should provide information about sleep; promote individualized motivation, intention, and planning around going to bed on time; and sensibly limit burden. However, neither intervention increased sleep duration or used a group format to positively shift attitudes or leverage social norms toward target goal engagement (25,31,38). Deeper emphasis, cued practice, and measurement regarding behavioral skills, such as sleep hygiene and management of unpleasant internal experiences might help to broaden and enhance benefits.

BP can indicate overmatched self-control (39), as it is often used as short-term distraction from negative emotion and cognition (36,40). Because this underregulated emotion and thought content interferes with one’s ability to prepare for sleep (e.g., 41–43), countermeasures should focus on improving self-regulation. Increasing self-compassion (SC) might facilitate effective emotion regulation, self-control, and sustained efforts to address BP by reducing maladaptive responses to setbacks and preserving motivation (e.g., 44–46).

Self-compassion enables individuals to step outside of distress-maintaining cognitive-emotional cycles, such as rumination and worry, and defuse them through decentered nonjudgmental, curious observation. Extending from this aware, stable mindset, self-compassion helps individuals neutralize set-back-related shame, guilt, and isolation by validating one’s connection to the shared human experience of pain, struggle, and imperfection. This connection to common humanity can then bolster the self-compassionate practice of rechanneling harsh self-criticism into soothing self-kindness. Also relevant to sleep, self-compassion is conceptualized as a means to switch from an overactive threat defense system, characterized by sympathetic nervous system (SNS) dominance, to engage the mammalian care system, characterized by equilibrating parasympathetic nervous (PNS) system activity (47). Distress is associated with SNS dominance and PNS suppression, whereas PNS activity is linked to safety, rest, healing, and growth. Research studying a range of populations indicates that greater practice of self-compassion is associated with increased mindfulness, self-compassion, emotion regulation, as well as lower stress, anxiety, depression, and emotional avoidance (48). Given that habitual bedtime procrastination and emotion dysregulation may be partly maintained by demotivating self-critical procrastinatory thinking (49), improving emotion regulation, procrastinatory cognition, and self-compassion might reduce BP most robustly (50). However, until now, no SC-focused intervention for BP has been developed or compared to a benchmark, such as sleep hygiene (SH) guidance.

### The Present Study

We have developed novel BP intervention structure and content meant to address perceived susceptibility, severity of consequences, benefits, barriers, behavioral intent and prompting from the HBM; attitudes, subjective norms, and perceived behavioral control from the TPB; and ambivalence resolution, planning, and public commitment from the TTM. We reasoned that a brief and accessible intervention might especially help individuals in busy contexts, where maintaining performance and motivation in the midst of setbacks is important. Therefore, this study recruited college students with BP and self-critical tendencies to examine the feasibility, acceptability, and preliminary efficacy of a multimodal intervention 1) involving a virtual group meeting and home practices, 2) leveraging models of positive health behavior change and maintenance, and 3) focusing on SC or SH. Based on pilot research of this intervention implemented in an in-person format with 25 individuals (51), we expected that both *virtual* interventions would be feasible and acceptable to participants (cf. 52,53 frameworks). We conceptualized that intervention focus on HBM, TPB, and TTM elements would manifest in assessments of post-meeting alliance and perceived group norms, which we hypothesized would positively predict engagement with home practices to reduce BP. We also predicted training-related BP and sleep insufficiency reductions, and moderated mediation whereby the relationship between increased SC and decreased BP would be mediated by improved emotion regulation for those with elevated reductions in procrastinatory cognition. Thus, based on a program outcomes logic model, we expected that bedtime procrastination and sleep would improve (ultimate outcomes) by way of training enhanced alliance, norms, and readiness for change (immediate outcomes), and with the support of lesson/practice engagement, self-compassion, sleep hygiene, emotion regulation, and procrastinatory cognition reduction (intermediate outcomes).

## Materials and Methods

### Participants

Ninety-three participants began the study, but 19 did not complete it (*n*^sleep hygiene^ = 12, 38.7%; *n*^self-compassion^ = 2, 8.3%; *n*^no group^ = 5, 13.2%). Participants more frequently discontinued if they had been assigned to the SH condition compared to the SC condition (*X*^2^ (1, *N* = 55) = 6.58, *p* = .01). The final sample included the 74 participants (*n*^sleep hygiene^ = 19, *n*^self-compassion^ = 22, *n*^no group^ = 33). There were no significant between-group differences among demographic variables (see Table 1) or prescreen sleep variables (see Supplemental Materials Table 1). Participants averaged 6-7 hours of sleep/night and reported high-average sleepiness; 28.41% endorsed above average sleepiness, i.e., > 10, on the Epworth Sleepiness Scale (54).

**Table 1.** Sample Demographic Characteristics

|  | *M or n* | *SD or %* |
| --- | --- | --- |
| Age | 25.68 | 7.67 |
| Persons Living in Home | 3.70 | 1.67 |
| Household Income (USD) | 70,315 | 67,477 |
| Gender |  |  |
| Cisgender woman | 66 | 89.2% |
| Cisgender man | 7 | 9.5% |
| Gender fluid | 1 | 1.4% |
| Race/Ethnicity |  |  |
| Black or African American | 5 | 6.8% |
| Latinx | 23 | 31.1% |
| White or Caucasian | 19 | 25.7% |
| Multiracial/Multiethnic | 8 | 10.8% |
| Other | 11 | 14.9% |
| Decline to Answer | 1 | 1.4% |
| Missing | 7 | 9.5% |
| Sexual Orientation |  |  |
| Bi-Sexual | 7 | 9.5% |
| Heterosexual | 52 | 70.3% |
| Lesbian | 1 | 1.4% |
| Other | 3 | 4.1% |
| Pansexual | 2 | 2.7% |
| Questioning | 1 | 1.4% |
| Decline to Answer | 1 | 1.4% |
| Missing | 7 | 9.5% |
| Relationship Status |  |  |
| Single | 30 | 40.5% |
| In a relationship | 20 | 27.0% |
| Married | 15 | 20.3% |
| Separated or Divorced | 2 | 2.7% |
| Other | 2 | 2.7% |
| Dating | 5 | 6.8% |
| Parental Status |  |  |
| Have child(ren) | 20 | 27.0% |
| Do Not Have Child(ren) | 54 | 73.0% |

### Procedure

Data were collected between September 2020 and May 2021, using the {*university*} research pool to recruit students prescreened for 1) dissatisfaction about BP, 2) self-critical tendencies, and 3) no significant insomnia (e.g., Insomnia Severity Index score < 15; Morin, 1993).

#### Inclusion Criteria and Surveys

We embedded several items within the Psychology Department mass testing prescreen survey. To screen for dissatisfaction about BP, we included the Bedtime Procrastination Scale item, “To what extent does this statement apply to you? I want to go to bed on time, but I just do not.” Participants responded using a five-point scale ranging from 1 (“Almost never”) to 5 (“almost always”). Those answering 2 (“sometimes”) or greater were recruited. To screen for possible procrastination-related self-critical tendencies, we included the Procrastinatory Cognitions Inventory item, “I’m letting myself down.” Participants responded using a five-point scale ranging from 0 (“Not at all”) to 4 (“All the time”). Those answering 1 (“sometimes”) or greater were recruited. Individuals who met initial criteria received email invitations or enrolled directly in the study online. Participants needed to understand English, provide consent, and agree to virtual group meeting rules.

Qualifying participants completed online Qualtrics surveys at baseline, immediately following the group training (if the participant was assigned to one), and post-intervention, i.e., after completion of the post-training self-practice portion of the intervention. At baseline, participants were first presented the informed consent document. At the end of it, they needed to answer two questions correctly to confirm comprehension of what they would be doing in the study before answering whether they consented to participate or not. Those who clicked “no” were exited from the survey and thus have no reportable data. For those who clicked “yes,” their non-identifiable response was attached to their survey data file, and they advanced within the survey to focal study measures. Specifically, participants completed self-report measures of bedtime procrastination, sleep hygiene, self-compassion, emotion regulation difficulties, procrastinatory cognition, readiness for change regarding sleep-enhancing behaviors, sleep insufficiency, mindfulness, positive and negative affect, self-control, and demographic information. Immediately following the meeting, participants privately answered questions regarding working alliance, subjective norms, and descriptive norms. At post-intervention, participants completed the same measures as at baseline, plus measures assessing relative acceptability of the intervention.

#### Group Trainings

Training conditions included one 1-hour virtual group meeting with between 3-6 individuals (*M* = 3.98; *SD =* 0.93). This format was chosen to maximize safety and accessibility around the COVID-19 pandemic onset, to minimize participant burden and potential for attrition, and to examine effects of a “minimal dose” of a group training intervention coupled with six home lessons/practices. Such an intervention format can be useful given that the modal number of behavioral health sessions people attend is one, and that college health centers and primary care behavioral health settings can deliver effective interventions at greater scale and efficiency than typical psychotherapy formats (55–57). Before leading groups, the clinical psychology graduate student co-leaders needed to demonstrate knowledge proficiency through an exam and skill proficiency through leading group simulations; post-simulation feedback was provided live and based on viewed video recordings by the PI/first author and by co-authors with more experience leading groups. To start meetings, co-leaders reviewed the meeting agenda with slides participants were emailed. Both group trainings followed semi-structured scripts with group expectation setting, educational content (how much sleep people need, sleep’s effects on health, and SH and BP paths to insufficient sleep), and prompts to elicit participant experiences and universalize salience and engagement norms. Each training also focused on SC or SH, as described below. At the end, key ideas were recapped, and to consolidate learning and elicit public commitment and planning, each member was asked, “what is one idea or practice you will take away from this meeting?” and “how will you apply it?”

Over the subsequent three weeks participants assigned to training conditions received six emailed home lessons/practices. These were framed as optional so that subsequent use would indicate participants’ intrinsically motivated engagement. All participants completed the final survey approximately 4 weeks after their first. For training completers, this survey assessed the training’s acceptability, home practice use frequency, and limited efficacy outcomes. Participants were debriefed and received course credit. Participants were identifiable by name to facilitate recruitment, retention, and course crediting. Thus, some authors had access to this information during participants’ study participation but not thereafter. Further, participants’ data were separated from any identifying information in the study’s aggregate data base, which included no identifying information. Procedures were approved by the [*university*] institutional review board.

##### Self-Compassion Training

The training drew heavily from the *Mindful Self-Compassion Workbook* (58), adapting material to fit the training format and connect it to BP. In the meeting, sections on “self-compassion components, misgivings, and realities,” components of SC, and the “mindful way through resistance and backdraft” were reviewed, and two brief mindfulness exercises were conducted (present awareness and the five senses, affectionate breathing). Media use as a potential form of “mindless” sleep-interfering BP behavior was also noted (12). Home lessons/practices included developing loving kindness, finding loving-kindness phrases, “finding your compassionate voice,” mindful compassionate preparation for bed, meeting difficult emotions, and self-appreciation.

##### Sleep Hygiene Training

The sleep hygiene training drew from <u>American Academy of</u> <u>Sleep Medicine</u>, <u>National Sleep Foundation</u>, and book materials (59,60), with the meeting including information, guidance, and discussion about circadian rhythm functioning, chronotype, sleep pressure, napping, sleep efficiency, stimulus control, optimal environmental conditions, physical activity, sleep-affecting substances, and pre-bedtime media use. Home lesson/practices focused on sleep promoting habits, wakefulness-promoting habits, managing sleep efficiency, creating an optimal sleeping environment, engaging physical activity conducive to sleep, and use/non-use of sleep-affecting substances.

### Measures

Unless otherwise specified, higher scores indicate higher levels of measured constructs below. Internal consistencies are listed in Tables.

#### Feasibility and Acceptability

A 14-item questionnaire regarding frequency of engagement with home exercises and other indicators of intervention acceptability based on Sekhon et al. (53). See Table 3 for items. Participants were also asked, “How many times did you use the following lesson/practices?” in reference to the six home practices they were emailed (scale 0 = none, 1= 1 time, 2 = 2-3 times, 3 = 4-6 times, 4 = 7 or more times.) Participants also provided qualitative feedback on the training by writing what they liked best and what they would change in a free-response text box.

#### Immediate Outcomes

##### Alliance and Norms

Working alliance and norms were assessed with a set of five items (see Table 2). The first three items, adapted from the Working Alliance Inventory-Short Revised (61), assessed for the three components of working alliance—bond, goals, and tasks. Items included, “This training and I have established a good understanding of the kind of changes that would be good for me.” (alliance goals); “I believe that the things I do from this training will help me to accomplish the changes that I want.” (alliance tasks); and “I believe the co-leaders I met today would accept, respect, and care about me, even if I did things that they do not approve of.” (alliance bond). The next two questions assessed for subjective norms and descriptive norms, based on the Theory of Planned Behavior (Fishbein & Ajzen, 1975). These items included, “I believe others in this group would be supportive if I adopted goals from the training” (TPB subjective norms) and “I believe others in this group will use the training’s strategies over the next 3 weeks.” (TPB descriptive norms). Participants were asked to consider to what extent each item statement corresponded with what their current beliefs, rating them on a scale ranging between 1 “does not correspond at all” and 7 (“corresponds exactly”) with a midpoint anchor of 4 (“corresponds moderately”). Higher scores indicate more positive alliance, attitudes, and beliefs.

**Table 2.**
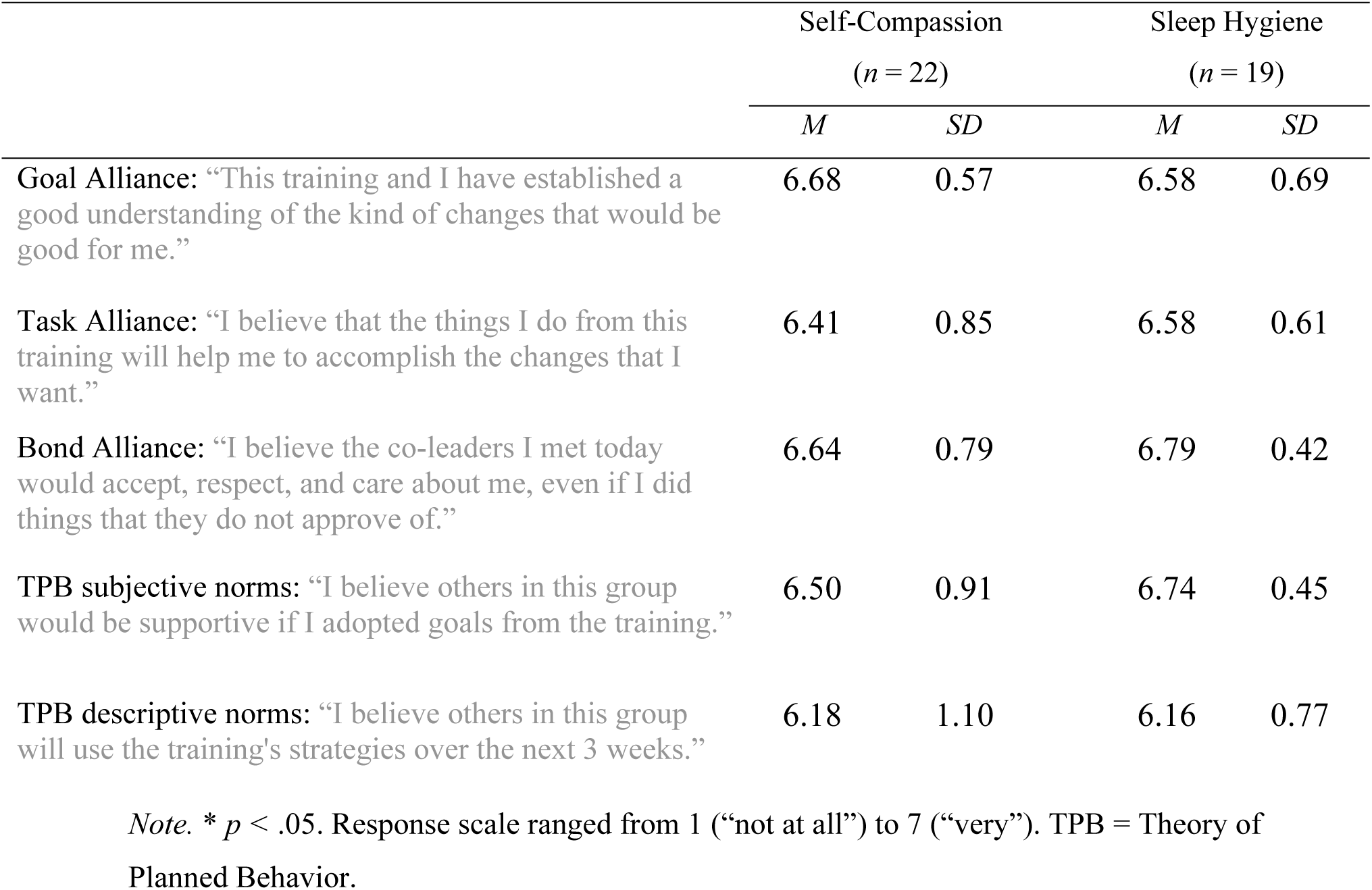
Indicators of Positive Alliance and Norms by Group Condition

##### Readiness for change

A single question was used to assess participants’ TTM stage of change regarding behaviors that might help them go to bed at night. They were asked, “Which of the following is the most accurate?” and then provided response choices ranging from 1 (precontemplation stage: “I have not been considering or trying behaviors that might help me go to bed at night in the last 6 months”) to 5 (maintenance stage: “I have been consistently implementing behaviors to help me go to bed at night for 3 months or more.”)

#### Intermediate Outcomes

##### Self-Compassion Scale-Short Form

The Self-Compassion Scale Short Form (SCS-SF; 62) is a 12-item self-report scale in which participants rate their self-compassionate or self-critical experiences on a 5-point scale ranging from “almost never” to “almost always.” The SCS-SF includes six self-compassionate phrases (“I try to see my failings as a part of the human condition,”) and six reverse-coded self-critical phrases (“I’m disapproving and judgmental about my own flaws and inadequacies”), with higher sum scores on the scale indicating higher levels of self-compassion. Total scores on the SCS-SF are closely correlated with total scores on the longer, 26-item Self-Compassion Scale (62).

##### Sleep Hygiene Scale

The Sleep Hygiene Scale (63) is a 19-item questionnaire designed to assess participants’ behaviors that facilitate or interfere with healthy sleep. The items on the scale describe various sleep-related behaviors as outlined by the International Classification of Sleep Disorders, Second Edition (ICSD-2), such as “Napped during the day,” and “Exercised within 4 hours of bedtime.” Two items reflect healthy sleep hygiene (“Woke up at approximately the same time; went to bed at approximately the same time”) and are reverse coded. The 19 items can be grouped into roughly five thematic groups – poor sleep scheduling, use of substances known to interfere with sleep quality (caffeine, alcohol, or tobacco), exhibition of sleep-disrupting activities, alternative uses for the bed, and sleep environment. Respondents were asked to, “Please indicate the average number of days per week in which you engaged in the following behaviors during the past month?” The 8-point response scale ranges from 0 (“0 days/week” to 7 (“7 days/week.”). Higher sum scores indicate worse overall SH.

##### Difficulties with Emotion Regulation Scale - Short Form

The Difficulties with Emotion Regulation Scale - Short Form (DERS-SF; 64) is an 18-item scale used to assess participants’ challenges with emotional clarity, nonacceptance of emotional responses, engaging in goal-directed behavior, impulse control, emotional awareness, and emotion regulation strategies. A sample item from the strategies subscale is “When I’m upset, it takes me a long time to feel better.” Items are all rated on a 5-point scale ranging between “Almost Never – 0-10%” to “Almost Always – 91-100%.” The DERS-18 has shown parallel structure and strong support for reliability and validity, similar to the original DERS (64).

##### Procrastinatory Cognitions Inventory

The Procrastinatory Cognitions Inventory (PCI; 65) is an 18-item questionnaire used to assess self-critical and fatalistic demotivating automatic thoughts associated with procrastination. PCI scores correlate with academic procrastination, brooding, perfectionism, depression, mindfulness, self-compassion (66,67). The PCI has been validated with college student populations (66,67) and correlates in expected directions with theoretically related constructs (e.g., academic procrastination, brooding rumination, performance avoidance, need for perfectionism, depressive symptoms, mindfulness, self-compassion). Response choices range from 0 (“not at all”) to (“all of the time”).

#### Ultimate Outcomes

##### Bedtime Procrastination Scale

The Bedtime Procrastination Scale is a validated 9-item self-report scale (9) that characterizes a participant’s habit of delaying bedtime not due to external reasons (i.e., due to circumstances within their control). Four of the items on the scale reflect effective self-control and are reverse-coded (“I can easily stop with my activities when it is time to go to bed,”) and five reflect the lack of control often exhibited in procrastinatory behavior (“I go to bed later than I had intended”). Response choices fall on a 5-point scale ranging from “almost never” to “almost always.” After the items are scored appropriately, the average score indicates the inclination of the participant to procrastinate at bedtime, with higher scores correlating with more bedtime procrastination. This measure was administered with instructions to answer in reference to last month.

##### Indicators of Sleep Insufficiency

Six items (8,9) were used to assess indicators of sleep insufficiency: experiencing insufficient sleep, going to bed late, daytime fatigue, (“In an average week, how many days do you feel you have had too little sleep?” “In an average week, how many days do you go to bed later than you would like to?” “In an average week, how many days do you feel tired during the day?”). Those questions were answered on 8-point scales ranging from “0 days” to “7 days”. Also, participants answered how problematic experiencing insufficient sleep, going to bed late, daytime fatigue was for them (e.g., “To what extent do you find it problematic if you go to bed later than you would like to?”) on a five-point scale ranging from 1 (“not at all”) to 5 (“very much.”)

#### Secondary Outcomes

##### Positive and Negative Affect Schedules

Participants completed a 10-item self-report measure modified version of the Positive and Negative Affect Schedules-Expanded Form (PANAS-X; 46,68) that assesses for affects most relevant to procrastination, six negative affect adjectives (i.e., distressed, upset, guilty, angry at self, dissatisfied with self, ashamed) and four positive affect adjectives (i.e., inspired, proud, thankful, hopeful). Participants were asked to read each adjective and rate “to what extent you have felt this way during the past 3-4 weeks” on a 5-point Likert-like scale ranging from 1 = “never” to 5 = “always.”

##### Five Facet Mindfulness Questionnaire 15

The Five Facet Mindfulness Questionnaire 15 (FFMQ-15; 69), distilled from the FFMQ-39, is a measure of participants’ attunement to five aspects of mindfulness: Observing, Describing, Acting with Awareness, Non-Judging of inner experience, and Non-Reactivity to inner experience. For example, one non-reactivity item reads, “When I have distressing thoughts or images, I am able just to notice them without reacting.” Participants rate the extent to which mindfulness-relevant statements describe them and their behavior on a 5-point scale ranging from 1 (“Never or very rarely true”) to 5 (“Very often or always true.”)

##### Brief Self-Control Scale

The Brief Self-Control Scale (BSCS; 70) is a commonly used 13-item measure focusing on individual differences in processes that involve self-control, such as breaking habits and working toward goals. For example, one item reads, “I am good at resisting temptations.” Items are rated on a 5-point scale ranging between 1 (“not at all”) and 5 (“very much).

#### Control Factor

##### External Reasons for Sleep Disturbance

The degree to which participants endorsed external reasons for sleep disturbances was assessed with the item, “To what extent is your bedtime generally affected by external circumstances that are outside of your control? Participants responded between 1 (Never) and 5 (Always). This item was administered at the beginning and end of the study. This measure was administered at the beginning and end of the study.

### Analytic Plan

Analyses were completed with SPSS Version 28.0 software. Means and standard deviations were calculated for all continuous variables, and correlational analysis and multiple regression analysis were used to examine relations of focal interest. Mixed-factorial ANOVA was used to examine group condition-by-time changes in variables of focal interest. Raw difference scores were used to evaluate change over time. Power analysis using G*Power indicated that a study comparing three conditions at two timepoints would have an 80% probability of detecting a medium sized true effect with a total sample of 66 participants. Based on large effects reported in previous studies of relevant BP interventions (36,37) and conservative prediction of at least medium-sized effects in the present study, we recruited a sample with adequate power. SPSS PROCESS model 14 (71) was used to test moderated mediation, specifically the hypothesis that mediation of the relation between self-compassion and BP by emotion regulation would be moderated by procrastinatory cognition. This model uses bootstrapping, a conventionally accepted data resampling technique, to estimate population parameters. Indirect effects’ reliability for evaluating moderated mediation are indicated when the range spanning the bootstrapped lower limit confidence interval (LLCI) and upper limit confidence interval (ULCI) excludes 0 (e.g., LLCI and ULCI are either both positive or both negative), based on 5,000 bootstrap samples. Emotion regulation and procrastinatory cognition scores were mean centered.

## Results

### Feasibility

#### Implementation, Practicality, and Adaptation

Co-leaders learned, practiced, and successfully implemented both trainings with the target population without unanticipated difficulties. Increasing directive elicitation of group interaction in training scripts was effective for the virtual meeting format.

#### Acceptability

Participant response statistics of measures of acceptability are shown in Table 3. Overall, *affective attitude* for both training groups was positive. When asked what they liked best, representative responses from SC participants included, “the consistency of the material,” “the meditation,” and “the variety of exercises.” SH participants liked the interpersonal connection and the *practicality* of the training (e.g., “simple to do but very effective.”) More detailed qualitative and quantitative responses are included in Supplemental Online Materials.

**Table 3.** Indicators of Acceptability by Group Condition

Indicators of Acceptability by Group Condition
|  | Self-Compassion<br>(*n* = 22) |  | Sleep Hygiene<br>(*n* = 19) |  |
| --- | --- | --- | --- | --- |
|  | *M* | *SD* | *M* | *SD* |
| ***Affective Attitude*** |  |  |  |  |
| How much did you enjoy the material and practices included in the full training (virtual group meeting + voluntary home lessons/practices)? | 6.05 | 1.21 | 5.61 | 1.50 |
| How valuable was the virtual group meeting? | 5.77 | 1.48 | 5.61 | 1.38 |
| How valuable were the home lessons/practices? | 5.40 | 1.47 | 5.72 | 1.32 |
| How likely would you be to utilize this material and these practices in the future? | 5.18 | 1.53 | 5.78 | 1.70 |
| ***Burden*** |  |  |  |  |
| For someone who struggles with getting to bed on time and related issues, how much does participating in this full training (virtual group meeting + voluntary home lessons/practices) get in the way of other values, needs, or commitments? | 3.67 | 2.01 | 2.83 | 1.72 |
| ***Intervention Coherence*** |  |  |  |  |
| To what extent did you understand how the lessons and practices were supposed to work? | 6.24 | 1.00 | 6.11 | 0.96 |
| ***Opportunity Cost*** |  |  |  |  |
| For someone who struggles with getting to bed on time and related issues, how worthwhile is the time spent in this training (virtual group meeting + voluntary lessons/practices)? | 5.95 | 1.05 | 5.89 | 1.23 |
| ***Perceived Effectiveness*** |  |  |  |  |
| To what extent does this training achieve its purpose (addressing struggles with getting to bed on time and related issues)? | 5.80 | 1.28 | 5.72 | 1.78 |
| ***Self-Efficacy*** |  |  |  |  |
| To what degree did you find this full training's lessons and practices easy to apply? | 5.52 | 1.69 | 5.50 | 1.25 |
| ***Ethicality*** |  |  |  |  |
| Considering your worldview and value system, how acceptable were the lessons and practices from this training? | 6.52 | 0.98 | 6.06 | 1.16 |

*Note.* \* $p < .05$ . Response scale ranged from 1 ("not at all") to 7 ("very"). Between-groups analyses were not conducted for the in-person modality given the minimal sample size. Categories and questions based on Sekhon et al. (2017) framework of acceptability of health interventions.

Regarding perceived *intervention coherence* and *self-efficacy*, ratings of the post-meeting “task” working alliance item and the post-training items assessing intervention coherence and self-efficacy suggested that, for both training conditions, participants understood the purpose of the material and practices, they found them easy to apply and relevant to their change goals. Participants rated the SC training as having less-than-moderate *burden*, slightly higher than the SH training vis-à-vis their other values, needs, or commitments. Many indicated they would not change anything about the training (SC: n=10 of 22; for SH: n=7 of 19), though, some suggested including more interactive check-ins and material support, such as on-demand online exercises and a sleep log.

Additionally, trainings were rated low in *opportunity cost* and high in *perceived effectiveness* and *ethicality,* with responses suggesting trainings were worthwhile and achieved their purpose. Participants highlighted the trainings’ unique value in comments (e.g., SC: “Helped me calm down during upsetting…times,” “it has improved my well-being quite a bit.”)

Regarding *demand*, all but one participant reported having engaged with at least some of the voluntary home practices from the trainings. On average SC participants used each practice 1-2 times, while SH participants used each practice 2-3 times.

### Preliminary Outcomes

Within- and between-condition differences were examined with 3 (condition: self-compassion, sleep hygiene, control) x 2 (time: pre- vs. post-training) mixed factorial ANOVAs.

#### Immediate outcomes

##### Alliance and Norms

Working alliance indicators (goals, tasks, and bond) and subjective and descriptive norms were all rated positively for both training conditions (Table 2). One-way ANOVA indicated a lack of differences between the self-compassion and sleep hygiene groups (all *p*’s > .05)

##### Readiness for Change

As anticipated, readiness for changing “behaviors to help (them) go to bed at night” increased only for the training conditions (see Table 4).

**Table 4.** Core Intermediate Training Variables

Core Intermediate Training Variables
| | Measure<br>Range | Study<br>$\alpha$ | Self-Compassion<br>($n = 22$) | | Sleep Hygiene<br>($n = 19$) | | No Group Control<br>($n = 32$) | |
| --- | --- | --- | --- | --- | --- | --- | --- | --- |
| | | | $M/d$ | $SD$ | $M/d$ | $SD$ | $M/d$ | $SD$ |
| BP T1 | 1-5 | .86 | 3.65<sup>a</sup> | 0.75 | 3.61<sup>b</sup> | 0.59 | 3.52 | 0.79 |
| BP T2 |  | .90 | 3.23<sup>a</sup> | 0.96 | 2.84<sup>b,c</sup> | 0.96 | 3.36<sup>c</sup> | 0.79 |
|  |  |  | *-0.57* |  | *- 0.94* |  | *-0.32* |  |
| Sleep Hygiene T1 | 0-133 | .71 | 48.64<sup>d</sup> | 16.97 | 43.63<sup>f</sup> | 17.31 | 48.82<sup>g</sup> | 15.95 |
| Sleep Hygiene T2 |  | .66 | 42.64<sup>d,e</sup> | 14.07 | 33.84<sup>e,f</sup> | 15.90 | 40.76<sup>g</sup> | 12.49 |
|  |  |  | *-0.65* |  | *-0.73* |  | *-0.76* |  |
| Self-compassion T1 | 5-60 | .82 | 34.68 | 9.41 | 33.74 | 9.10 | 35.45 | 7.26 |
| Self-compassion T2 |  | .84 | 35.82 | 10.60 | 36.84 | 8.15 | 36.15 | 6.36 |
|  |  |  | *0.20* |  | *0.27* |  | *0.14* |  |
| DERS T1 | 18-90 | .87 | 40.91 | 12.67 | 40.42 | 15.82 | 40.11 | 9.75 |
| DERS T2 |  | .89 | 40.50 | 15.39 | 39.58 | 12.03 | 40.73 | 10.29 |
|  |  |  | *-0.04* |  | *-0.07* |  | *0.08* |  |
| PCI T1 | 0-76 | .92 | 36.73 | 16.66 | 36.84<sup>h</sup> | 12.49 | 31.61 | 11.95 |
| PCI T2 |  | .93 | 34.14 | 16.53 | 29.00<sup>h</sup> | 12.18 | 31.39 | 13.46 |
|  |  |  | *-0.26* |  | *-0.59* |  | *-0.03* |  |
| Readiness T1 | 1-5 | N/A | 1.95<sup>i</sup> | 1.33 | 2.21<sup>j</sup> | 1.36 | 2.08 | 1.32 |
| Readiness T2 |  |  | 3.23<sup>i</sup> | 0.97 | 3.42<sup>j,k</sup> | 2.35 | 1.35<sup>k</sup> | 1.34 |
|  |  |  | *0.86* |  | *0.48* |  | *-0.66* |  |

Note. \* $p < .05$ . Superscript letters represent paired contrasts that are significantly different at $p < .05$ alpha. For instance, the <sup>c</sup> pair represents the significant *increase* in mean days from time 1 to time

#### Secondary Outcomes: Affect, Mindfulness, and Self-Control

As shown in Table 6, negative affect decreased for both training groups. Positive affect, mindfulness, and self-control all increased in the SH condition only.

**Table 6.**
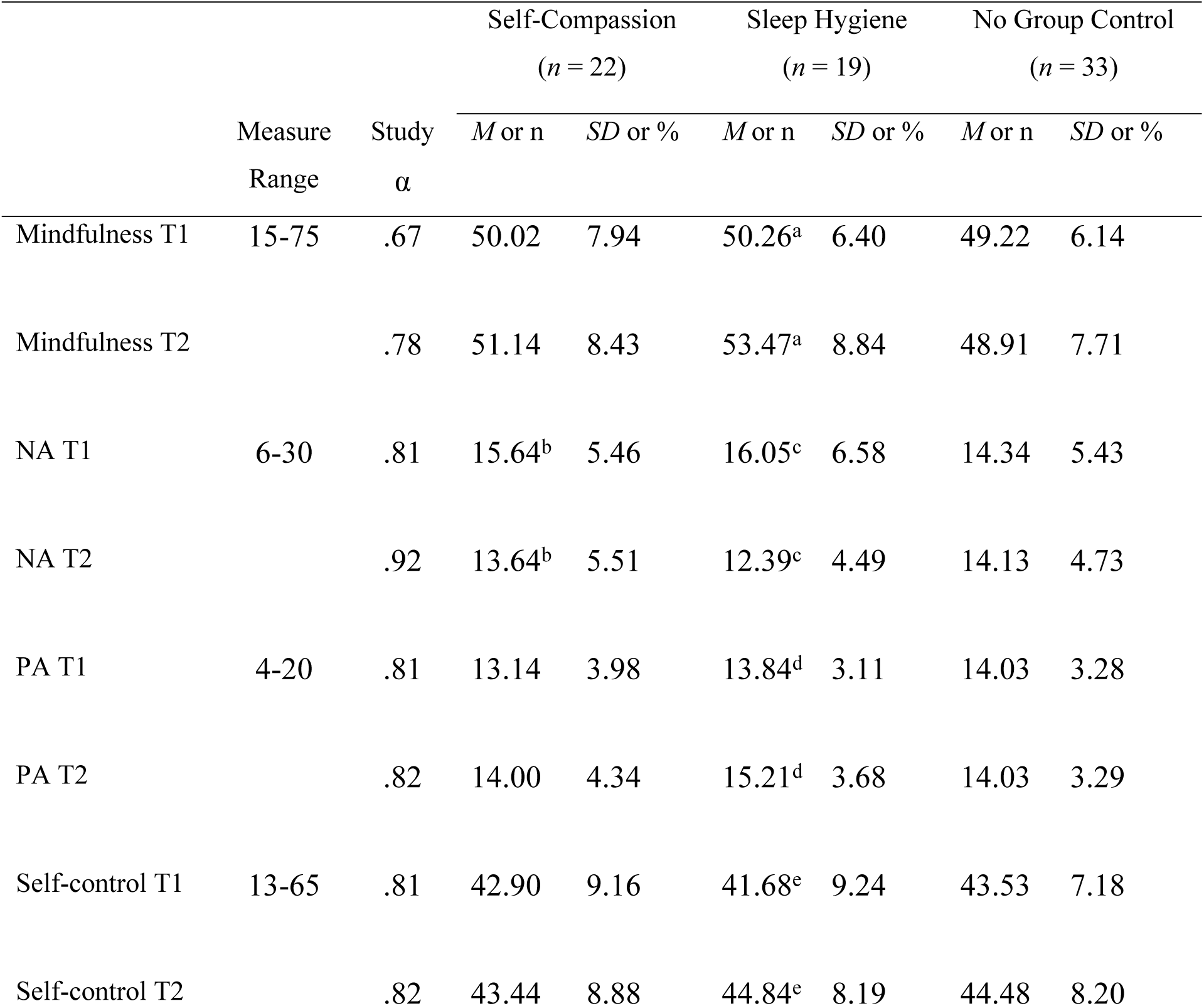
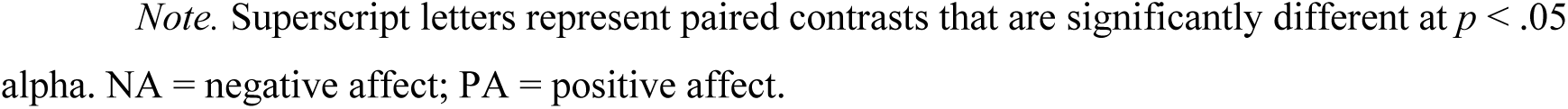
Measures of Possible Sequelae

| | Measure<br>Range | Study<br>$\alpha$ | Self-Compassion<br>(*n* = 22) | | Sleep Hygiene<br>(*n* = 19) | | No Group Control<br>(*n* = 33) | |
| --- | --- | --- | --- | --- | --- | --- | --- | --- |
|  |  |  | *M* or *n* | *SD* or % | *M* or *n* | *SD* or % | *M* or *n* | *SD* or % |
| Mindfulness T1 | 15-75 | .67 | 50.02 | 7.94 | 50.26<sup>a</sup> | 6.40 | 49.22 | 6.14 |
| Mindfulness T2 |  | .78 | 51.14 | 8.43 | 53.47<sup>a</sup> | 8.84 | 48.91 | 7.71 |
| NA T1 | 6-30 | .81 | 15.64<sup>b</sup> | 5.46 | 16.05<sup>c</sup> | 6.58 | 14.34 | 5.43 |
| NA T2 |  | .92 | 13.64<sup>b</sup> | 5.51 | 12.39<sup>c</sup> | 4.49 | 14.13 | 4.73 |
| PA T1 | 4-20 | .81 | 13.14 | 3.98 | 13.84<sup>d</sup> | 3.11 | 14.03 | 3.28 |
| PA T2 |  | .82 | 14.00 | 4.34 | 15.21<sup>d</sup> | 3.68 | 14.03 | 3.29 |
| Self-control T1 | 13-65 | .81 | 42.90 | 9.16 | 41.68<sup>e</sup> | 9.24 | 43.53 | 7.18 |
| Self-control T2 |  | .82 | 43.44 | 8.88 | 44.84<sup>e</sup> | 8.19 | 44.48 | 8.20 |

*Note.* Superscript letters represent paired contrasts that are significantly different at $p < .05$ alpha. NA = negative affect; PA = positive affect.

## Discussion

This study found evidence supporting the feasibility and acceptability of brief virtual group-plus-self-directed self-compassion (SC) and sleep hygiene (SH) trainings that incorporated core elements of the health belief model, theory of planned action, and transtheoretical model of behavior change. Trainings were adapted to promote fluid, respectful, meaningful, and confidential interactions in a video conferencing format. This format minimized burden, increased reach, and addressed the recommendation from initially piloting an in-person version of the trainings (51). Participant feedback indicated that the format and training material worked well, and that the novel SC training provided valued knowledge and experiences not included in existing BP interventions.

As hypothesized, home practice use was strongly predicted by health behavior model-relevant factors, such goal-related alliance, task-related alliance, readiness for change, and subjective norms that were targeted in the intervention. On the other hand, the implied within-group variability in these predictors underscores the need for intervention to individually address salience, confusion, and skepticism for every group member early on. However, it is noteworthy that both interventions increased participants’ readiness to change BP behavior with medium-to-large effects, while the no-intervention control did not.

This study also found preliminary evidence of efficacy and possible mechanisms of change. There were medium-to-large effect size training-related improvements with BP, sleep insufficiency, and daytime fatigue across both trainings. Moreover, improved BP strongly related to improved sleep outcomes, including sleep insufficiency—contrasting with previously published BP-interventions. Additionally, the hypothesized moderated mediation model was supported.

The fact that the relation between increased SC and decreased BP was mediated by improved emotion regulation for only those with the largest reductions in procrastinatory cognition prompts several important considerations. First, trainings did not quantifiably improve emotion regulation or SC at group levels (though both measures did not anchor participant ratings to the previous month as BP and SH measures did). It is possible that boosting specific intervention emphasis on self-compassion, emotion regulation, and their interrelation might strengthen not only self-compassion and emotion regulation effects, but also downstream effects on BP and sleep. Second, it can only be speculated about whether stronger intervention effects on SC would have led to group level increases in SC or a significant direct effect of SC on BP. However, the *non*significant direct effect of increasing self-compassion on decreasing BP found in this study suggests that increasing SC alone was insufficient to change BP over time; SC worked through intermediate mechanisms, such as effectively regulating emotion *and* procrastinatory cognition. Third, the lack of a significant indirect effect between SC and BP through emotion regulation for those with low or medium reductions in procrastinatory cognition suggests that our intervention (and perhaps others) might be improved by increasing emphasis on how individuals can use self-compassion to defuse patterns of thinking that interfere with goal attainment, such as going to bed on time. Fourth, model outcomes suggest that for many, decreases in BP may come through SH improvements. However, although SH behavior may be easier to boost than trait SC or emotion regulation, improving SC may be an important orthogonal path to improve emotion regulation, unhelpful thinking patterns, and BP for some (72). Extending on prior cross-sectional research (Sirois et al., 2019), boosting SC was associated with negative affect decreases in this study.

Participant suggestions to increase impact, practicality, and reach of the trainings focused on providing access to more materials and more interactive encounters. Increasing the “dose” of this study’s SC intervention in these ways might lead to a stronger and broader effect on individuals’ SC and emotion regulation, given that greater home practice was related to SC increases (cf. 73). Greater practice could have helped individuals become increasingly aware of self-criticism’s effects and increasingly flexible in handling challenges and setbacks with acceptance, self-kindness, self-protection, self-motivation, and perceived connection to human experience (Neff, 74,75). For example, daytime fatigue decreased in both trainings, but only SC participants endorsed daytime fatigue as being less problematic, suggesting they also shifted how they relate to daily challenges.

A combined-integrated SC and SH training could potentially synthesize benefits and address limitations of each approach (cf. 76). For example, the SH condition uniquely improved mindfulness, positive affect, self-control, and procrastinatory cognition suggests that this approach should be included. Implementing and observing concrete SH changes could initiate sleep improvements that predispose more salutary cycles of cognitive, affective, and behavioral regulation, and bolstering the more abstract, incremental work toward effecting lasting changes in self-compassion. Meanwhile, the SC condition’s lower dropout rate, compared to the SH condition may indicate greater personal salience, enthusiasm, and engagement, which could help sustain integrated SC/SH groups. SC requires emotional vulnerability, and having peers reciprocating vulnerability facilitates safety, trust, modeling, and hope (77). Emotion regulation and SH are important pathways to support healthier sleep patterns (40), and people need self-regulatory intervention options that give them autonomy and flexibility (37).

### Strengths and Limitations

Regarding study strengths, this study addresses the need for intervention research on BP by extending beyond existing interventions, evaluating quantitative and qualitative participant feedback, and examining both preliminary efficacy and possible pathways of change. Key study limitations focus on methodological design, content, level of analysis, and population focus. We employed the most commonly used measure of BP but did not track daily BP and sleep in a way that could counteract retrospective recall inaccuracies (78). Relatedly, our design precluded assessing longevity, bidirectionality, or nonlinear temporal dynamics of effects. Factors examined are likely embedded within biopsychosocial regulatory systems. Thus, an extension of the present research could examine psychophysiological correlates of sleep-wake cycle factors and thoroughly assess behavioral indicators of sleep health, chronotype, and medical conditions that could affect pre-bedtime behavior (also see 79–81). For example, some research suggests that mindfulness and self-compassion practices may increase levels of oxytocin, which can support better mood regulation and sleep (82,83).

Additionally, this study’s sample was predominantly within the young adult age range, so it is unknown to what degree that study findings would generalize to younger or older populations given life-stage-related biological and psychosocial factors can impact bedtime behaviors and sleep significantly (84,85). Also, although simulated groups were recorded and evaluated for intervention fidelity during training, actual group meetings were not recorded to maximize perceived safety and acceptability for participants, and thus fidelity was not evaluated. Thus, future studies could examine effects of developmental life stage and intervention fidelity. Another useful extension of the present research would be to examine history of psychological trauma given that trauma exposure is associated with low SC, emotion dysregulation, and sleep difficulties (86).

Lastly, our trainings’ virtual meeting format took on increased poignancy during the COVID-19 pandemic. Although distress, anxiety, and social disconnection were not explicitly measured in the present study, one might assume that average levels of these variables were elevated based on anecdotal conversations with participants and generalizable phenomena during the period (84). Future studies should assess these variables and adapt interventions for clinical health settings to examine how well results from this college student sample generalize.

### Conclusions

There is a need to learn more about the mechanisms involved in BP and to integrate and refine mechanism-focused interventions that are acceptable and effective with diverse populations. The present study represents a step in that direction. It can help support and strengthen important connections among immediate outcomes (alliance, norms, readiness for change), intermediate outcomes (lesson/practice engagement, self-compassion, sleep hygiene, emotion regulation, and procrastinatory cognition), and ultimate outcomes (bedtime procrastination, sleep insufficiency) as a cohesive, comprehensive framework for effectively and efficiently addressing bedtime procrastination. Needed future studies should include larger samples, judiciously expanded trainings, broader assessment of health effects, and sequencing that maximizes efficacy and participant engagement.

## Data Availability

Study data can be accessed via https://osf.io/sbn67.

## Acknowledgments

The authors would like to acknowledge the contributions of Larissa Farah, M.A., Staci Grant, Psy.D. and Kristina Harper, Psy.D. in performing literature searches and contributing to discussions that deepened the lab focus on the potential of mindfulness and self-compassion within interventions. We also valued the expert consultation of Mindful Self-Compassion trainer, Gwen Brehm, M.Ed., LPC, LMFT of the Center for Mind-Body Medicine. Lastly, we would like to acknowledge and thank Lauren Palmer and Harleen Sandhu, M.D., for contributions in helping this research project function. The authors received no specific funding for this work.

## Declaration of Interest Statement

The authors have no financial or non-financial interests to disclose.

## Notes

### Competing Interest Statement

The authors have declared no competing interest.

### Author Declarations

This research was conducted under the approval of the Committee for the Protection of Human Subjects institutional review board at the University of Houston-Clear Lake (approval date: 9/2/2020) and in accordance with the Declaration of Helsinki.

## References

1. Ram S, Seirawan H, Kumar SKS, Clark GT. Prevalence and impact of sleep disorders and sleep habits in the United States. Sleep Breath. 2010 Feb;14(1):63–70.

2. Blask DE. Melatonin, sleep disturbance and cancer risk. Sleep Med Rev. 2009 Aug;13(4):257–64.

3. Buxton OM, Marcelli E. Short and long sleep are positively associated with obesity, diabetes, hypertension, and cardiovascular disease among adults in the United States. Soc Sci Med. 2010 Sep;71(5):1027–36.

4. Irwin MR, Olmstead R, Carroll JE. Sleep Disturbance, Sleep Duration, and Inflammation: A Systematic Review and Meta-Analysis of Cohort Studies and Experimental Sleep Deprivation. Biol Psychiatry. 2016 Jul;80(1):40–52.

5. Blaszczynski A, Gordon K, Silove D, Sloane D, Hillman K, Panasetis P. Psychiatric morbidity following motor vehicle accidents: A review of methodological issues. Compr Psychiatry. 1998 May;39(3):111–21.

6. Kalsi J, Tervo T, Bachour A, Partinen M. Sleep versus non−sleep-related fatal road accidents. Sleep Med. 2018 Nov;51:148–52.

7. Irish LA, Kline CE, Gunn HE, Buysse DJ, Hall MH. The role of sleep hygiene in promoting public health: A review of empirical evidence. Sleep Med Rev. 2015 Aug;22:23–36.

8. Kadzikowska-Wrzosek R. Self-regulation and bedtime procrastination: The role of self-regulation skills and chronotype. Personal Individ Differ. 2018 Jul;128:10–5.

9. Kroese FM, De Ridder DTD, Evers C, Adriaanse MA. Bedtime procrastination: introducing a new area of procrastination. Front Psychol. 2014;5:611.

10. Kroese FM, Evers C, Adriaanse MA, de Ridder DT. Bedtime procrastination: A self-regulation perspective on sleep insufficiency in the general population. J Health Psychol. 2016 May;21(5):853–62.

11. Nauts S, Kamphorst BA, Stut W, De Ridder DTD, Anderson JH. The Explanations People Give for Going to Bed Late: A Qualitative Study of the Varieties of Bedtime Procrastination. Behav Sleep Med. 2019 Nov 2;17(6):753–62.

12. Chung SJ, An H, Suh S. What do people do before going to bed? A study of bedtime procrastination using time use surveys. Sleep [Internet]. 2020 Apr 15 [cited 2021 Oct 20];43(4). Available from: https://doi.org/10.1093/sleep/zsz267

13. Hill VM, Rebar AL, Ferguson SA, Shriane AE, Vincent GE. Go to bed! A systematic review and meta-analysis of bedtime procrastination correlates and sleep outcomes. Sleep Med Rev. 2022 Dec;66:101697.

14. Herzog-Krzywoszanska R, Krzywoszanski L. Bedtime Procrastination, Sleep-Related Behaviors, and Demographic Factors in an Online Survey on a Polish Sample. Front Neurosci. 2019 Sep 18;13:963.

15. Kim G, Jeon H, Suh S. Bedtime Procrastination as a Mediator in the Relationship Between Active Emotion Regulation Strategies and Insomnia. J Sleep Med. 2021 Dec 31;18(3):175– 81.

16. You Z, Mei W, Ye N, Zhang L, Andrasik F. Mediating effects of rumination and bedtime procrastination on the relationship between Internet addiction and poor sleep quality. J Behav Addict. 2020 Dec 31;9(4):1002–10.

17. Hu W, Ye Z, Zhang Z. Off-Time Work-Related Smartphone Use and Bedtime Procrastination of Public Employees: A Cross-Cultural Study. Front Psychol. 2022 Mar 10;13:850802.

18. Rubin R. Matters of the Mind—Bedtime Procrastination, Relaxation-Induced Anxiety, Lonely Tweeters. JAMA. 2020 Jan 7;323(1):15.

19. Armitage CJ, Conner M. Efficacy of the Theory of Planned Behaviour: A meta-analytic review. Br J Soc Psychol. 2001 Dec;40(4):471–99.

20. Badawi J, Bistricky SL, Millmann M, Gimenez-Zapiola M, Pascuzzi B, Short MB. Brief online intervention model promotes sustained helping behavior across 6 months following a population-wide traumatic event. Psychol Rep. in press;

21. Bistricky SL, Harper KL, Roberts CM, Cook DM, Schield SL, Bui J, et al. Understanding and Promoting Stress Management Practices Among College Students Through an Integrated Health Behavior Model. Am J Health Educ. 2018 Jan 2;49(1):12–27.

22. Carpenter CJ. A Meta-Analysis of the Effectiveness of Health Belief Model Variables in Predicting Behavior. Health Commun. 2010 Nov 30;25(8):661–9.

23. Harper K, Short MB, Bistricky S, Kusters IS. 1-2-3! Catch-Up for HPV: A Theoretically Informed Pilot Intervention to Increase HPV Vaccine Uptake among Young Adults. Am J Health Educ. 2023 Mar 4;54(2):119–34.

24. Harrison JA, Mullen PD, Green LW. A meta-analysis of studies of the Health Belief Model with adults. Health Educ Res. 1992;7(1):107–16.

25. McEachan RRC, Conner M, Taylor NJ, Lawton RJ. Prospective prediction of health-related behaviours with the Theory of Planned Behaviour: a meta-analysis. Health Psychol Rev. 2011 Sep;5(2):97–144.

26. Mead MP, Irish LA. Application of health behaviour theory to sleep health improvement. J Sleep Res [Internet]. 2020 Oct [cited 2023 Mar 25];29(5). Available from: https://onlinelibrary.wiley.com/doi/10.1111/jsr.12950

27. Thomas MC, Duggan KA, Kamarck TW, Wright AGC, Muldoon MF, Manuck SB. Conscientiousness and Cardiometabolic Risk: A Test of the Health Behavior Model of Personality Using Structural Equation Modeling. Ann Behav Med. 2022 Jan 1;56(1):100–11.

28. Champion VL, Skinner CS. The health belief model. In: Health behavior and health education: Theory, research, and practice, 4th ed. San Francisco, CA, US: Jossey-Bass; 2008. p. 45–65.

29. Fry JP, Neff RA. Periodic Prompts and Reminders in Health Promotion and Health Behavior Interventions: Systematic Review. J Med Internet Res. 2009 May 14;11(2):e16.

30. Rosenstock IM. Historical Origins of the Health Belief Model. Health Educ Monogr. 1974 Dec;2(4):328–35.

31. Fishbein M, Ajzen I. Belief, attitude, intention, and behavior: an introduction to theory and research. Reading, Mass: Addison-Wesley Pub. Co; 1975. 578 p. (Addison-Wesley series in social psychology).

32. Kor K, Mullan BA. Sleep hygiene behaviours: An application of the theory of planned behaviour and the investigation of perceived autonomy support, past behaviour and response inhibition. Psychol Health. 2011 Sep;26(9):1208–24.

33. McCaul KD, Hinsz VB, McCaul HS. The Effects of Commitment to Performance Goals on Effort1. J Appl Soc Psychol. 1987 May;17(5):437–52.

34. Miller WR, Rose GS. Toward a theory of motivational interviewing. Am Psychol. 2009;64(6):527–37.

35. Grandner MA. Obstacles to overcome when improving sleep health at a societal level. In: Sleep and Health [Internet]. Elsevier; 2019 [cited 2023 Mar 26]. p. 107–15. Available from: https://linkinghub.elsevier.com/retrieve/pii/B9780128153734000095

36. Suh S, Cho N, Jeoung S, An H. Developing a Psychological Intervention for Decreasing Bedtime Procrastination: The BED-PRO Study. Behav Sleep Med. 2022 Nov 2;20(6):659– 73.

37. Valshtein TJ, Oettingen G, Gollwitzer PM. Using mental contrasting with implementation intentions to reduce bedtime procrastination: two randomised trials. Psychol Health. 2020 Mar 3;35(3):275–301.

38. Sheeran P, Orbell S. Augmenting the Theory of Planned Behavior: Roles for Anticipated Regret and Descriptive Norms1. J Appl Soc Psychol. 1999 Oct;29(10):2107–42.

39. Kamphorst BA, Nauts S, De Ridder DTD, Anderson JH. Too Depleted to Turn In: The Relevance of End-of-the-Day Resource Depletion for Reducing Bedtime Procrastination. Front Psychol. 2018 Mar 14;9:252.

40. Lin SY, Chung KKH. Chronotype and trait self-control as unique predictors of sleep quality in Chinese adults: The mediating effects of sleep hygiene habits and bedtime media use. Sirois FM, editor. PLOS ONE. 2022 Apr 15;17(4):e0266874.

41. Campbell RL, Bridges AJ. Bedtime procrastination mediates the relation between anxiety and sleep problems. J Clin Psychol. 2022 Sep 28;jclp.23440.

42. Teoh AN, Ooi EYE, Chan AY. Boredom affects sleep quality: The serial mediation effect of inattention and bedtime procrastination. Personal Individ Differ. 2021 Mar;171:110460.

43. Zhang C, Meng D, Zhu L, Ma X, Guo J, Fu Y, et al. The Effect of Trait Anxiety on Bedtime Procrastination: the Mediating Role of Self-Control. Int J Behav Med [Internet]. 2022 Apr 22 [cited 2022 Dec 29]; Available from: https://link.springer.com/10.1007/s12529-022-10089-3

44. Abdoli N, Farnia V, Radmehr F, Alikhani M, Moradinazar M, Khodamoradi M, et al. The effect of self-compassion training on craving and self-efficacy in female patients with methamphetamine dependence: a one-year follow-up. J Subst Use. 2021 Sep 3;26(5):491–6.

45. Biber DD, Ellis R. The effect of self-compassion on the self-regulation of health behaviors: A systematic review. J Health Psychol. 2019 Dec;24(14):2060–71.

46. Sirois FM, Nauts S, Molnar DS. Self-Compassion and Bedtime Procrastination: an Emotion Regulation Perspective. Mindfulness. 2019 Mar;10(3):434–45.

47. Gilbert P. The origins and nature of compassion focused therapy. Br J Clin Psychol. 2014 Mar;53(1):6–41.

48. Neff KD. Self-Compassion: Theory, Method, Research, and Intervention. Annu Rev Psychol. 2023 Jan 4;74(1):annurev-psych-032420-031047.

49. Sass SM, Early LM, Long L, Burke A, Gwinn D, Miller P. A brief mindfulness intervention reduces depression, increases nonjudgment, and speeds processing of emotional and neutral stimuli. Ment Health Prev. 2019 Mar;13:58–67.

50. Deng Y, Ye B, Yang Q. COVID-19 Related Emotional Stress and Bedtime Procrastination Among College Students in China: A Moderated Mediation Model. Nat Sci Sleep. 2022 Aug;Volume 14:1437–47.

51. Bistricky SL, Lopez AK, Pollard TB, Egan A, Brehm G. The rationale, feasibility, and acceptability of a brief intervention addressing bedtime procrastination and sleep through self-compassion and sleep hygiene. Unpublished.

52. Bowen DJ, Kreuter M, Spring B, Cofta-Woerpel L, Linnan L, Weiner D, et al. How We Design Feasibility Studies. Am J Prev Med. 2009 May;36(5):452–7.

53. Sekhon M, Cartwright M, Francis JJ. Acceptability of healthcare interventions: an overview of reviews and development of a theoretical framework. BMC Health Serv Res. 2017 Dec;17(1):88.

54. Johns MW. A New Method for Measuring Daytime Sleepiness: The Epworth Sleepiness Scale. Sleep. 1991 Nov 1;14(6):540–5.

55. Phillips J, Salsman NL, Rigdon D, Taylor J, Buell J, Ronis-Tobin V. Implementation of an integrated primary care behavioral health training model on campus. J Am Coll Health. 2022 Nov 10;1–4.

56. Reiter JT, Dobmeyer AC, Hunter CL. The Primary Care Behavioral Health (PCBH) Model: An Overview and Operational Definition. J Clin Psychol Med Settings. 2018 Jun;25(2):109– 26.

57. Young J, Dryden W. Single-session therapy – past and future: an interview. Br J Guid Couns. 2019 Sep 3;47(5):645–54.

58. Neff KD, Germer CK. The mindful self-compassion workbook: a proven way to accept yourself, build inner strength, and thrive. New York, NY: Guilford Press; 2018. 206 p.

59. Jacobs GD. Say good night to insomnia. Updated ed. New York: Henry Holt; 2009. 211 p. (A Holt paperback).

60. Walker MP. Why we sleep: unlocking the power of sleep and dreams. First Scribner hardcover edition. New York: Scribner, an imprint of Simon & Schuster, Inc; 2017. 360 p.

61. Hatcher RL, Gillaspy JA. Development and validation of a revised short version of the working alliance inventory. Psychother Res. 2006 Jan;16(1):12–25.

62. Raes F, Pommier E, Neff KD, Van Gucht D. Construction and factorial validation of a short form of the Self-Compassion Scale. Clin Psychol Psychother. 2011 May;18(3):250–5.

63. Gellis LA, Lichstein KL. Sleep Hygiene Practices of Good and Poor Sleepers in the United States: An Internet-Based Study⋆⋆This work was supported by a grant from the University of Memphis, Department of Psychology. Behav Ther. 2009 Mar;40(1):1–9.

64. Victor SE, Klonsky ED. Validation of a Brief Version of the Difficulties in Emotion Regulation Scale (DERS-18) in Five Samples. J Psychopathol Behav Assess. 2016 Dec;38(4):582–9.

65. Stainton M, Lay, CH, Flett GL. Trait procrastinators and behavior/trait-specific cognitions. J Soc Behav Personal. 2000;15(5):297–312.

66. Flett AL, Haghbin M, Pychyl TA. Procrastination and Depression from a Cognitive Perspective: An Exploration of the Associations Among Procrastinatory Automatic Thoughts, Rumination, and Mindfulness. J Ration-Emotive Cogn-Behav Ther. 2016 Sep;34(3):169–86.

67. Flett GL, Stainton M, Hewitt PL, Sherry SB, Lay C. Procrastination Automatic Thoughts as a Personality Construct: An Analysis of the Procrastinatory Cognitions Inventory. J Ration-Emotive Cogn-Behav Ther. 2012 Dec;30(4):223–36.

68. Watson D, Clark LA, Tellegen A. Development and validation of brief measures of positive and negative affect: The PANAS scales. J Pers Soc Psychol. 1988;54(6):1063–70.

69. Baer RA, Carmody J, Hunsinger M. Weekly Change in Mindfulness and Perceived Stress in a Mindfulness-Based Stress Reduction Program: Weekly Change in Mindfulness and Stress in MBSR. J Clin Psychol. 2012 Jul;68(7):755–65.

70. Tangney JP, Baumeister RF, Boone AL. High Self-Control Predicts Good Adjustment, Less Pathology, Better Grades, and Interpersonal Success. J Pers. 2004;72(2):271–324.

71. Hayes AF. Introduction to mediation, moderation, and conditional process analysis: a regression-based approach. Third edition. New York; London: The Guilford Press; 2022. 731 p. (Methodology in the social sciences).

72. Teixeira I, Simões S, Marques M, Espírito-Santo H, Lemos L. Self-criticism and self-compassion role in the occurrence of insomnia on college students. Eur Psychiatry. 2016 Mar;33(S1):s268–s268.

73. Smeets E, Neff K, Alberts H, Peters M. Meeting Suffering with Kindness: Effects of a Brief Self-Compassion Intervention for Female College Students: Self-Compassion Intervention for Students. J Clin Psychol. 2014 Sep;70(9):794–807.

74. Neff KD. Fierce self-compassion: how women can harness kindness to speak up, claim their power, and thrive. First edition. New York, NY: HarperWave, an imprint of HarperCollins Publishers; 2021. 374 p.

75. Wilson AC, Mackintosh K, Power K, Chan SWY. Effectiveness of Self-Compassion Related Therapies: a Systematic Review and Meta-analysis. Mindfulness. 2019 Jun;10(6):979–95.

76. Resick PA, Galovski TE, Uhlmansiek MO, Scher CD, Clum GA, Young-Xu Y. A randomized clinical trial to dismantle components of cognitive processing therapy for posttraumatic stress disorder in female victims of interpersonal violence. J Consult Clin Psychol. 2008 Apr;76(2):243–58.

77. Yalom ID, Leszcz M. The theory and practice of group psychotherapy. Sixth edition. New York: Basic Books; 2020.

78. Sobell LC, Sobell MB. Timeline Follow-Back. In: Litten RZ, Allen JP, editors. Measuring Alcohol Consumption [Internet]. Totowa, NJ: Humana Press; 1992 [cited 2022 Dec 29]. p. 41–72. Available from: http://link.springer.com/10.1007/978-1-4612-0357-5_3

79. Chen D, Zhang Y, Lin J, Pang D, Cheng D, Si D. Factors influencing bedtime procrastination in junior college nursing students: a cross-sectional study. BMC Nurs. 2022 Dec;21(1):97.

80. Exelmans L, Van den Bulck J. Self-control depletion and sleep duration: the mediating role of television viewing. Psychol Health. 2018 Oct 3;33(10):1251–68.

81. Martin M, Sadlo G, Stew G. The phenomenon of boredom. Qual Res Psychol. 2006 Jan;3(3):193–211.

82. Bellosta-Batalla M, Blanco-Gandía M, Rodríguez-Arias M, Cebolla A, Pérez-Blasco J, Moya-Albiol L. Brief mindfulness session improves mood and increases salivary oxytocin in psychology students. Stress Health. 2020 Oct;36(4):469–77.

83. Lipschitz DL, Kuhn R, Kinney AY, Grewen K, Donaldson GW, Nakamura Y. An Exploratory Study of the Effects of Mind–Body Interventions Targeting Sleep on Salivary Oxytocin Levels in Cancer Survivors. Integr Cancer Ther. 2015 Jul;14(4):366–80.

84. Foster R. Life Time: The New Science of the Body Clock, and How It Can Revolutionize Your Sleep and Health. London, UK: Penguin; 2022.

85. Roberts CM, Harper KL, Bistricky SL, Short MB. Bedtime behaviors: Parental mental health, parental sleep, parental accommodation, and children’s sleep disturbance. Child Health Care. 2020 Apr 2;49(2):115–33.

86. Scoglio AAJ, Rudat DA, Garvert D, Jarmolowski M, Jackson C, Herman JL. Self-Compassion and Responses to Trauma: The Role of Emotion Regulation. J Interpers Violence. 2018 Jul;33(13):2016–36.

